# Short-term Outcomes in Children Recovered from Multisystem Inflammatory Syndrome associated with SARS-CoV-2 infection

**DOI:** 10.1101/2021.06.23.21259292

**Authors:** Sibabratta Patnaik, Mukesh Kumar Jain, Sakir Ahmed, Arun Kumar Dash, P Ram Kumar, Bandya Sahoo, Reshmi Mishra, Manas Ranjan Behera

**Affiliations:** Kalinga Institute of Medical Sciences, Bhubaneswar, India; Dept of Clinical Immunology and Rheumatology, Kalinga Institute of Medical Sciences, Bhubaneswar, India

**Author notes:** **Correspondence to:** Sibabratta Patnaik, Associate Professor, Paediatrics, Kalinga Institute of Medical Sciences, Bhubaneswar, India, Mail.

**Keywords:** follow-up, MIS-C, multi-inflammatory syndrome, outcomes, paediatric COVID-19

## Abstract

**Background:** Multi System Inflammatory Syndrome in children (MIS-C) associated with COVID-19 is a recently recognised potentially life-threatening entity. There is limited data on post MIS-C sequelae.

**Methods:** 21 children fulfilling the WHO criteria for MIS-C were included in our study. Data was collected at baseline and at 12-16 weeks post discharge to look for any persistent sequelae mainly relating to the lungs or heart including coronary arteries

**Results:** Fever was the most common presentation, found in 18 (85.7%) patients. All had marked hyper-inflammatory state. Low ejection fraction (EF) was found in 10 (47.6%), but none had any coronary artery abnormality. All received corticosteroids, while 7 (33.3%) children required additional treatment with intravenous Immunoglobulins. 20 children improved while 1 left against medical advice. At discharge, 3 children had impaired left ventricular function. At median 15 weeks’ follow-up, no persistent complications were found. EF had returned to normal and no coronary artery abnormalities were found during repeat echocardiography. Chest radiographs showed no fibrosis and all biochemical parameters had normalized.

**Conclusion:** The children with MIS-C are extremely sick during the acute stage. Timely and adequate management led to full recovery without any sequelae at a median follow-up of 15 weeks.

## Introduction

Multi-System Inflammatory Syndrome in children associated with COVID 19 (MIS-C) is a relatively new problem precipitated by the coronavirus pandemic. Since first time reported from England in April 2020[1], this disease has emerged as a problem for children and adolescents. It was shown to present with acute heart failure as a part of an hyper-inflammatory response[2]. As the disease is relatively new, many new concepts are coming up in recent studies. MIS-C has a broad spectrum of presentation, some having mild symptoms while others have severe symptoms requiring intensive care support[3]. The disease usually presents 6 to 8 weeks after the viral infection and thought to be immune mediated. The hyper-inflammation in MIS-C differs from that of acute COVID-19 [4]. The MIS-C cases have many similarities with Kawasaki disease (KD), though MIS-C children are substantially older and have much severe inflammation. A quarter of patients have features of KD, while another quarter of patients may have features of incomplete KD[5]. Though the disease presents with profound inflammation, the mortality is around 1-2%[5, 6].

However, there is little knowledge about the squeal or post-discharge complications of this set of patients. Like KD, some may have pre-mature coronary artery disease with acute cardiac event. The SARS-CoV-2 virus activates the thrombosis cascade via several mechanisms and also induces the overexpression of plasminogen activator inhibitor□1 (PAI□1) [7]. PAI-1 has also been found to be overexpressed in Kawasaki disease and is associated with development of coronary artery aneurysms [8]. Thus, patients with MIS-C may have a risk of developing coronary artery aneurysms on follow-up.

Similar illness can also be caused by other respiratory viruses or emerging new viral illnesses. Thus there is an urgent need to follow up a cohort of MIS-C patients to determine their short term and long term outcomes. We have examined the short term outcomes in our cohort of 21 children with MIS-C including detailed clinical and laboratory data.

## Methods

### Participants

A cohort of MIS-C was built at the department of Paediatrics of Kalinga Institute of Medical Sciences, Bhubaneswar over the last 1 year. All the children below 19 years fitting to WHO criteria[9] for MIS-C were included in the study. This included fever, rash, shock, heart failure, coagulopathy, gastrointestinal manifestations in the presence raised acute phase reactants, evidence of recent COVID-19 infection and exclusion of other causes of sepsis. For all patients RT-PCR testing of nasopharyngeal swab was carried out for SARS-CoV-2. Antibodies to SARS-COV-2 were detected using Elecsys® immunoassay kit [Roche diagnostics, Basel, Switzerland] as per the manufacturer’s instructions. The assay uses recombinant protein representing the nucleocapsid antigen for detecting the antibody.

### Data collection

Data collected at admission, included age, gender, clinical profile, investigations including echocardiography and chest radiographs. During hospital stay, treatments provided and outcomes till discharge were recorded.

### Follow-up

After discharge, children were followed-up monthly for persistent clinical features or the emergence of any new symptoms. At 3 to 4 months after discharge, patients were routinely investigated for complete blood count, C-reactive protein (CRP), renal function, liver function, D-dimer, ferritin and lactate dehydrogenase (LDH). Chest radiographs and echocardiography was done for all patients.

### Statistics

Statistical analysis was carried out with SPSS software version 20. Normality of data were checked by the Shapiro Wilk test. Since the data were non-parametric, continuous variables were expressed as median, with inter-quartile range. Categorical variables were expressed as frequencies (percentages). Mann-Whitney U was used to compare means of continuous variables while the Fisher Exact test was used to compare proportions of categorical variables. P value less than 0.05 was taken as statistically significant.

### Ethics

Ethical clearance had been obtained from the Institute Ethics Committee of the Kalinga Institute of Medical Sciences, Bhubaneswar (Reference no: KIIT/KIMS/IEC/542/2021) for this study. Written informed consent was taken from parents.

## Result

There were a total 21 children enrolled in our cohort; 13(62%) of them were male, with mean age of presentation was 8.48 (± 4.3) years. Fever was the most common manifestation followed by rash. The children had features of involvement of the gastrointestinal system, the respiratory system and the cardiovascular system [Table 1].

**Table I:**
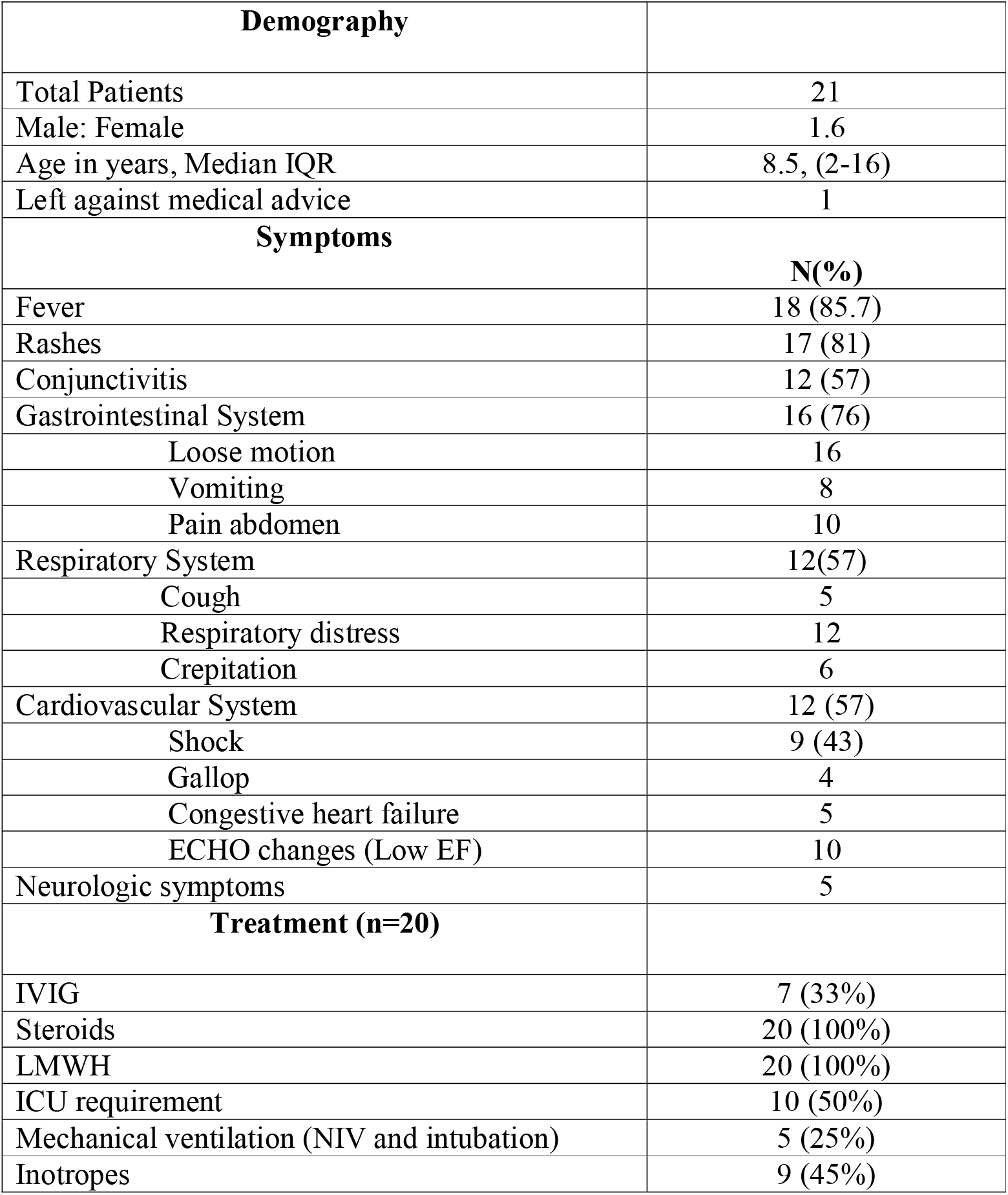
Demography and Clinical Characteristics.

| **Demography** |  |
| --- | --- |
| Total Patients | 21 |
| Male: Female | 1.6 |
| Age in years, Median IQR | 8.5, (2-16) |
| Left against medical advice | 1 |
| **Symptoms** | **N(%)** |
| Fever | 18 (85.7) |
| Rashes | 17 (81) |
| Conjunctivitis | 12 (57) |
| Gastrointestinal System | 16 (76) |
| Loose motion | 16 |
| Vomiting | 8 |
| Pain abdomen | 10 |
| Respiratory System | 12(57) |
| Cough | 5 |
| Respiratory distress | 12 |
| Crepitation | 6 |
| Cardiovascular System | 12 (57) |
| Shock | 9 (43) |
| Gallop | 4 |
| Congestive heart failure | 5 |
| ECHO changes (Low EF) | 10 |
| Neurologic symptoms | 5 |
| **Treatment (n=20)** |  |
| IVIG | 7 (33%) |
| Steroids | 20 (100%) |
| LMWH | 20 (100%) |
| ICU requirement | 10 (50%) |
| Mechanical ventilation (NIV and intubation) | 5 (25%) |
| Inotropes | 9 (45%) |

Nine (43%) children developed shock, requiring critical care with inotropic support, either at presentation or during hospital stay. Only one child had features of encephalitis. Twenty (95%) children had high titre of COVID-19 antibody. Two children were positive by both RTPCR and antibody testing. Only a few of the parents (28.6%) could remember contact with COVID-19 patients in past 6-8 weeks.

There was marked hyper-inflammatory state as evident by various acute phase reactants [Table II]. The majority of patients had normal leucocyte counts. Anaemia and hypoalbuminemia were common. On echocardiography,10 children had low ejection fraction. However, none of the children had coronary artery abnormality.

**Table II:**
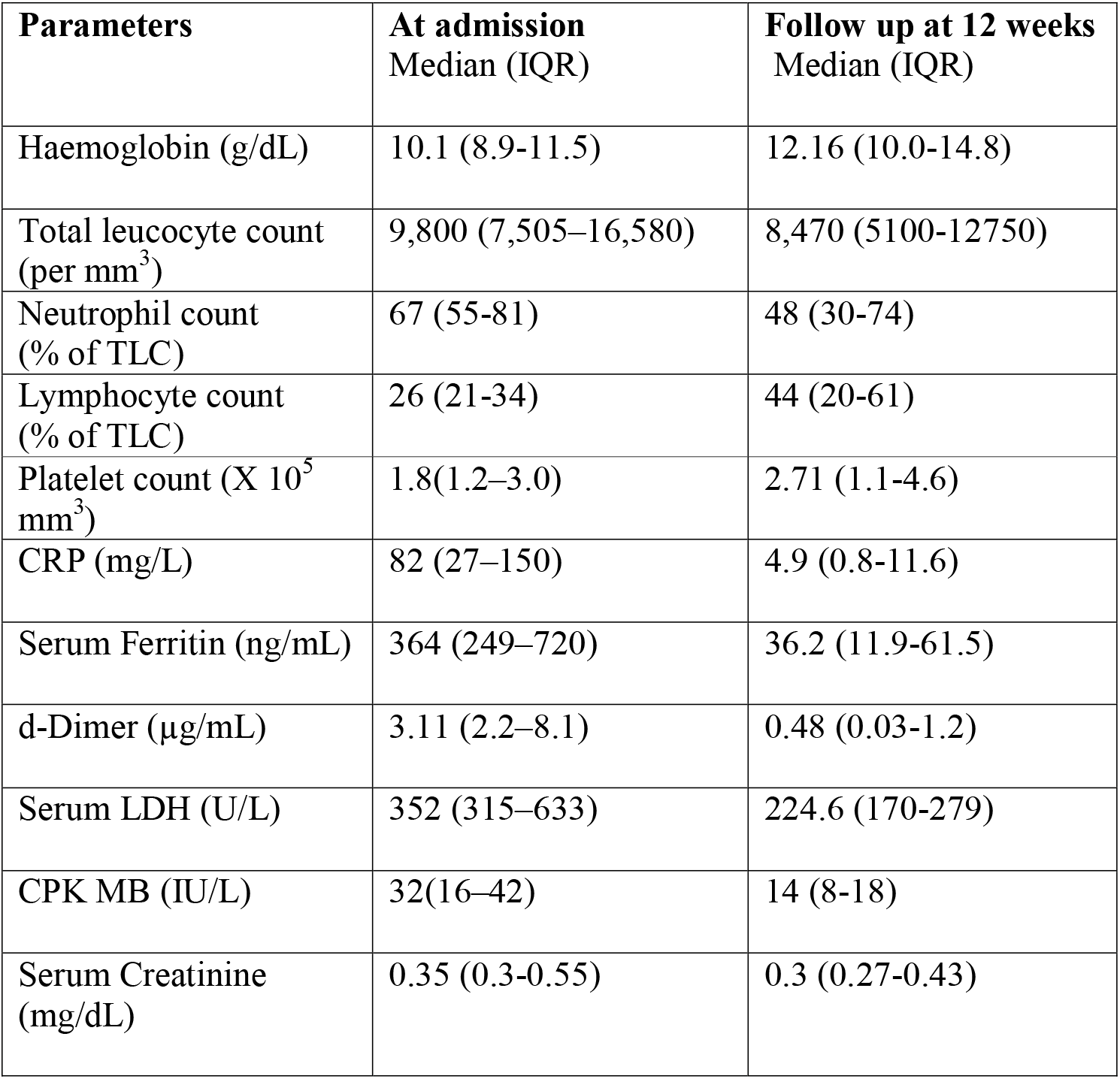
Laboratory Profile of children with MIS-C.

### Outcome at discharge

Out of 21 children, 1 child left against medical advice while 20 children continued treatment in our hospital. Ten (50%) were managed in the paediatric intensive care unit with indications of either respiratory distress needing oxygen support, shock requiring inotropes or congestive cardiac failure. Five (25%) children required non-invasive ventilation (NIV) support and only 1 child was intubated and ventilated. One child with mild symptoms was treated with oral steroid while rest 19 (95%) children received intravenous methyl-prednisolone (MPS). Intravenous immunoglobulin (IVIG) was used along with intravenous MPS in 7 (35%) patients. Post pulse MPS for 3 days, patients received tapering doses of oral steroids over the next 2-3 weeks. Low molecular weight heparin (LMWH) was introduced to all cases as per protocol. The average stay in the hospital was 8.6 (± 3.2) days. At discharge; all were hemodynamically stable, while 3 (15%) had mild left ventricular (LV) dysfunction.

### Follow-up at 3-4 months

Between 12 to 16 weeks after discharge, 16 children turned up for follow up. All children were hemodynamically stable without any major health problem. We repeated the blood counts and biochemistry. The hematologic and inflammatory parameters had normalised in all patients. (Table II) Follow up echocardiography of 15 children were normal, there were no evidence of coronary dilatation in any of our patients. One child who had global hypokinesia at admission, was found to have mild tricuspid regurgitation at 12 weeks’ follow-up. Another girl had feature of astasia abasia. She presented with inability to walk properly though clinical examination revealed no abnormality. All relevant investigations including magnetic resonance imaging of the brain and electroencephalogram were normal. The symptoms subsided in 12 weeks with proper counselling and psychiatrist consultation.

## Discussion

Our follow-up of MIS-C patients was to determine any sequel or delayed manifestation of this newly-described syndrome. In our cohort, we did not find any such sequel including any development of coronary artery aneurysms.

At presentation, three patients did not have any history of fever. Though by definition, fever was an essential criterion, they matched all other criteria for MIS-C [9].They might have had mild to moderate fever at onset which was missed at home. Reviews of previous cases have also revealed fever in around 80% cases [10].

In a pooled meta-analysis of features of MIS-C, myocardial involvement was approximately found in 60% [11]. This is comparable with that of our cohort. Initial reports of COVID-19 associated MIS-C cases describe ventricular dysfunction, coronary artery changes, atrioventricular valve regurgitation and pericardial effusions [2, 12, 13]. This warranted proper follow up of these cases to look for long term sequel. In our study we could show that none of these re-occurred in the follow-up period of 3-4 months.

We found neurologic involvement in four children, and one of them presented with features of encephalitis. A review that included 6 cohorts of MIS-C, found that 38% out of 187 children had some neurological issues [14]. Post-infectious immune mediated mechanism might be responsible for such issues, though more studies are required to substantiate it. Similar to myocardial involvement, there have been several reports of brain involvement in MIS-C and serial imaging of both the brain and the heart have been suggested [15].

Psychological issue in the form of astasia-abasia was found in one adolescent girl, few days after discharge from hospital. It was attributed to a somatoform disorder induced by ICU stay. She had improved over a period of 12 weeks.

Gastrointestinal involvement was common (76%) in our cohort. There are case reports of MIS-C patients presenting with pain abdomen and diarrhoea, and then developing Kawasaki like features including mucositis, and coronary artery dilatation [16]. Some patients have been admitted with a suspicion of appendicitis [17].

Maculopapular rashes and conjunctival congestion can be considered as features overlapping with complete and incomplete Kawasaki disease; and has been reported in 30-70% cases [11]. High d-dimer is found to be associated with poor outcome in many studies, but exact implication of high D-dimer is not yet understood [18].

As MIS-C is a hyper-inflammatory condition; immunomodulators play a major role in management. All of our patients had received corticosteroids. The decision to start IVIG in 7 patients was taken based on severity of the condition and also if the patient had not improved after the initial methyl prednisolone pulse. Combination of IVIG and methyl prednisolone is recommended for high risk cases including infants and in patients with coronary changes [19].

The American College of Rheumatology has recommended therapeutic anticoagulation in cases with ejection fraction (EF) <35%, documented thrombosis or coronary artery z-score more than 10 [19]. All of our patients had received LMWH. Severe disease is more common in 6-12year age group and the children with increased concentrations of CRP, ferritin, D-dimer and low platelets and lymphocytes were more likely to have shock or require ICU admission [20].

Supplemental oxygen was required in 10 patients; 5 required NIV support while a child with features of cardiogenic shock with pulmonary oedema required intubation and ventilation. A similar study from Mumbai in India had demonstrated high rate of invasive ventilation (39.1%) [21]. In the previously mentioned systematic review, the need for invasive ventilation was around 25%, while vasopressors were needed in more than 50% [3].

All the children were asymptomatic and clinically stable during follow up between 12-16 weeks after discharge. Laboratory parameters were within normal limits. Three children had mild LV dysfunction at discharge that normalized on follow-up. One study on echocardiographic follow-up of MIS-C patients for 7 days has shown good recovery of cardiac function [22].

In a recent Indian study of 32 patients, ventricular dysfunction was found in 13 patients; 11 had normal ejection fraction at 2 weeks follow up while rest 2 were also normal by 6 weeks [16]. Persistent abnormalities in strain and diastolic function in patients with MIS-C with normal EF has also been reported [17]. This points to requirement of multicentre longitudinal follow up studies for MIS-C.

Neurological manifestations improved before discharge in all, but one adolescent girl had somatoform disorder which persisted for more than 8 weeks. Amongst COVID-19 survivors, 17.4% had anxiety disorder, and 13.7% had mood disorder at 6-month follow-up [23]. Severe disease is associated with more chance of psychological issues.

There was no mortality reported in our cohort. In a systematic review, the mortality was described 18 (1.9%) deaths out of 953 cases [5]. Most of these cases presented with shock, and many had comorbidities like G6PD deficiency, obesity, leukaemia, asthma and chronic neurologic disorders. In a multi-centric cohort of 76 cases of MIS-C, 75 (98.6%) cases recovered without a significant sequel [24].

The limitation of our study include the small number of patients. However, MIS-C is a comparatively rare complication and to the best of our knowledge, limited cases of follow-up of such patients after discharge from hospital has been reported till now [24, 25].

## Conclusion

MIS-C is a new challenge in this COVID-19 pandemic era. A Broad case definition, lack of specific diagnostic tests and management guidelines, and uncertainty about long-term outcomes are major hurdles in its management. Our cohort showed that the morbidity of MIS-C was reversible with proper treatment and there was no significant complication or sequel at a median of 3.5-month follow-up. This study helps to dispel concerns of development of Kawasaki-like coronary artery aneurysms in MIS-C patients, if timely treatment is given. Longer studies with larger number of patients can confirm this.

## Data Availability

AVAILABLE

## Abbreviations

MIS-C: Multi System Inflammatory Syndrome in children
EF: Ejection fraction
KD: Kawasaki Disease
PAI-1: plasminogen activator inhibitor□1
CRP: C-reactive protein
LDH: Lactate dehydrogenase
LMWH: Low molecular weight heparin

